# GWAS of Depression Phenotypes in the Million Veteran Program and Meta-analysis in More than 1.2 Million Participants Yields 178 Independent Risk Loci

**DOI:** 10.1101/2020.05.18.20100685

**Authors:** Daniel F. Levey, Murray B. Stein, Frank R. Wendt, Gita A. Pathak, Hang Zhou, Mihaela Aslan, Rachel Quaden, Kelly M. Harrington, Gerard Sanacora, Andrew M. McIntosh, John Concato, Renato Polimanti, Joel Gelernter, on behalf of the Million Veteran Program

## Abstract

We report a large meta-analysis of depression using data from the Million Veteran Program (MVP), 23andMe Inc., UK Biobank, and FinnGen; including individuals of European ancestry (n=1,154,267; 340,591 cases) and African ancestry (n=59,600; 25,843 cases). We identified 223 and 233 independent SNPs associated with depression in European ancestry and transancestral analysis, respectively. Genetic correlations within the MVP cohort across electronic health records diagnosis, survey self-report of diagnosis, and a 2-item depression screen exceeded 0.81. Using transcriptome-wide association study (TWAS) we found significant associations for gene expression in several brain regions, including hypothalamus (NEGR1, p=3.19×10^−25^) and nucleus accumbens (DRD2, p=1.87×10^−20^). 178 genomic risk loci were fine-mapped to find likely causal variants. We identified likely pathogenicity in these variants and overlapping gene expression for 17 genes from our TWAS, including TRAF3. This study sheds light on the genetic architecture of depression and provides new insight into the interrelatedness of complex psychiatric traits.

## INTRODUCTION

Depression is the most common mental health condition, with lifetime prevalence in the U.S. of more than 20%^1^. Over 300 million people, or 4.4% of the world’s population, are estimated to be affected by depression, which imposes substantial costs on individuals and on society at large. Health expenditures exceeded $90 billion for treatment of depression and anxiety disorders in the U.S. in 2013.^2^ There also is a substantial personal cost to depression; for example, 60% of people who die by suicide have a diagnosed mood disorder. Indeed, depression and mood disorders have been shown to have genetic overlap with suicidal behavior in several recent studies.^3–6^

Only recently has substantial progress has been made in understanding the underlying genetic architecture of depression, led by the Psychiatric Genomics Consortium (PGC) and a large meta-analysis combining results from PGC,^7^ UKB,^8^ FinnGen (http://r2.finngen.fi/pheno/F5_MOOD) and 23andMe.^9,10^ In this article, we describe genome-wide association analysis of ∼310,000 participants from the U.S. Department of Veterans Affairs (VA) Million Veteran Program (MVP). MVP is one of the largest and most diverse biobanks in the world with genetic and electronic health record data available. When combined with the prior analysis from PGC, UK Biobank, and 23andMe,^10^ over one million participants were available for this study, the largest genetic analysis of depression to date.

We identified 178 genetic risk loci and 223 independently significant SNPs. We used the summary statistics from this analysis to investigate genetic correlations between depression, and several cohorts with different phenotypic assessments as well as overlap with other related traits. We used genomic SEM to examine shared genetic architecture and pleiotropy among complex traits. We also investigated functional consequences through fine mapping analysis, transcriptomic enrichment with respect to multiple brain tissues, and functional annotation. The results provide a deep look into the genetic architecture of depression and the underlying complex biology.

## Results

### Primary analysis

For the International Classification of Diseases (ICD) code definition of depression (hereafter, “ICD-Depression”) (see Online Methods for detailed diagnosis definitions), the phenotype with the most available data for the MVP cohort, we conducted a GWAS on 250,215 European individuals (83,810 cases). These MVP data were then included in a meta-analysis in METAL using inverse variance weighting with available depression GWAS summary statistics from three large cohorts of European-ancestry subjects (Figure 1a, Table 1): the PGC and the UK Biobank,^10^ FinnGen (http://r2.finngen.fi/pheno/F5_MOOD), and 23andMe,^9^ for a total of 1,154,267 subjects of European ancestry (330,173 cases). We identified 223 independent significant SNPs at 178 genomic risk loci in the primary analysis of European ancestry (Figure 1). We also conducted a GWAS in the African American (AA) sample from MVP in 59,600 participants (25,843 cases). There were no GWS findings from our primary analysis of ICD-Depression in African American ancestry, so examined overlap with the 223 GWS SNPs from our primary ICD-Depression meta-analysis of European American ancestry. Of the 223 GWS SNPs from the primary analysis, 206 were available following QC in the AA cohort. 61% (n=125) of the EUR SNPs had the same direction of effect in AAs, with 20 nominally significant (p<0.05) and 1 surviving Bonferroni correction (Figure 5). Finally, we conducted a transancestral meta-analysis by combining the results from the primary GWAS from European and African ancestry. This transancestral analysis of 366,434 cases and 847,433 controls identified 233 independent significant SNPs at 183 genomic risk loci (Figure 5).

**Figure 1.**
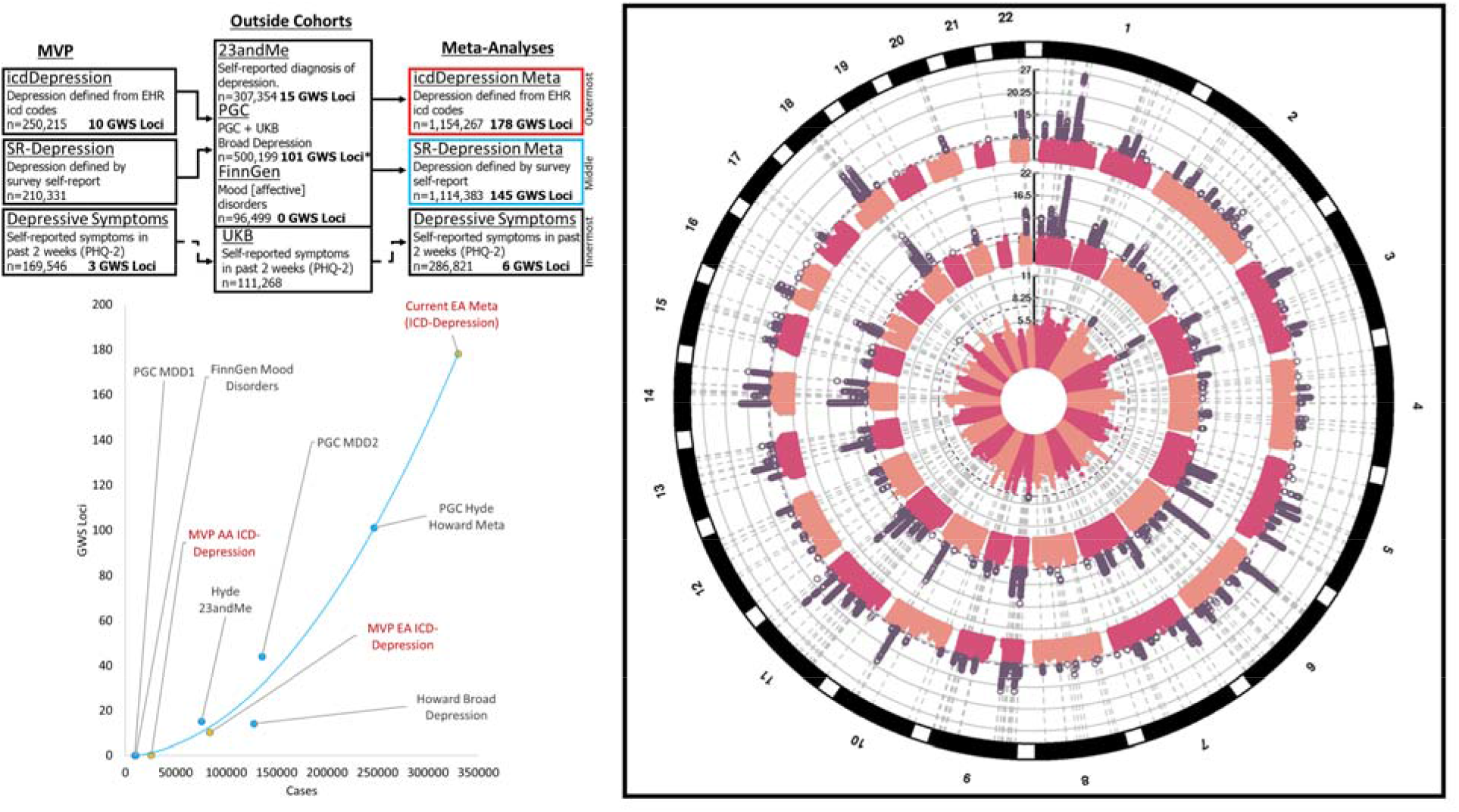
**Design of the study and circular Manhattan Plot.** Left Panel: Design of the study (top). Three phenotypes were evaluated within MVP: ICD-Depression (innermost ring, right panel) which was derived from ICD codes, SR-Depression (middle ring, right panel) which was defined by self-reported diagnosis of depression in the MVP survey, and Depressive symptoms (outermost ring, right panel) which come from the PHQ2 2-item scale found in the MVP survey. ICD-Depression and SR-Depression were each meta-analyzed with depression results from:23andMe, PGC, and FinnGen. MVP PHQ2 was meta-analyzed with results from the PHQ2 2-item scale from UK biobank. Right Panel: Circular Manhattan Plot. Significant results are highlighted in purple. Lower left Panel: Accelerating pace of loci discovery in depression GWAS. Y axis indicates the number of discovered loci in a study, with the X axis showing the number of cases included in each study. Red text and yellow markers indicate original analyses conducted for this study using MVP data for EA, AA and the overall ICD-Depression meta-analysis of EAs.

**Table 1.** Demographics.

| *Cohort* | *Case* | *Control* | *Total* |
| --- | --- | --- | --- |
| MVP ICD-Depression | 83,810 | 166,405 | 250,215 |
| MVP SR-Depression | 55,228 | 155,103 | 210,331 |
| 23andMe self-reported diagnosis of depression | 75,607 | 231,747 | 307,354 |
| UKB/PGC PGC + UKB Broad Depression | 170,756 | 329,443 | 500,199 |
| FinnGen Mood [affective] disorders | 10,418 | 86,081 | 96,499 |
| **ICD-Depression Meta**<br>**(MVP ICD-Depression + 23andMe +**<br>**UKB/PGC + FinnGen)** | **340,591** | **813,676** | **1,154,267** |
| **SR-Depression Meta**<br>**(MVP SR-Depression + 23andMe +**<br>**UKB/PGC + FinnGen)** | **312,009** | **802,374** | **1,114,383** |
| MVP PHQ2 |  |  | 175,553 |
| UKB PHQ2 |  |  | 111,268 |
| **PHQ2 Meta**<br>**(MVP PHQ2 + UKB PHQ2)** |  |  | **286,821** |

**Table 2.** Progress and History of Depression GWAS.

| Study | Cases | GWS loci |
| --- | --- | --- |
| PGC MDD1 | 9,240 | 0 |
| Howard Broad Depression | 127,552 | 14 |
| Hyde 23andMe | 75,607 | 15 |
| PGC MDD2 | 135,458 | 44 |
| PGC Hyde Howard Meta | 246,363 | 101 |
| FinnGen Mood Disorders | 10,418 | 0 |
| **MVP ICD-Depression** | **83,810** | **10** |
| **MVP African ancestry ICD-Depression** | **25,843** | **0** |
| **Current Meta (ICD-Depression)** | **340,591** | **178** |

### Secondary phenotype definitions

A similar meta-analysis was conducted using self-reported (SR)-Depression (see Methods) from MVP, conducted on 210,331 individuals who completed survey items on self-reported diagnosis of depression by a medical professional; the total meta-analysis with the traits from PGC, UK Biobank and FinnGen included 1,114,383 subjects. A third analysis considered depressive symptoms in the past two weeks from the Patient Health Questionniare-2 (PHQ-2),^11^ a 2-item scale which assesses depressive symptoms within the past two weeks (Table S3). For this phenotype, data were only available from MVP and UK Biobank, with a total sample of 286,821 European participants.

### Linkage Disequilibrium Score Regression (LDSC)

LDSC was used in two ways: 1) to identify genetic correlations and SNP-based heritability within each of the depression cohorts and phenotypes; and 2) to identify genetic correlation with other traits based on the primary meta-analysis (ICD-Depression). Heritability in the primary ICD-Depression meta-analysis was 11.3% (z=29.63, sample prevalence 28.6%, population prevalence 20%), while heritability in the secondary analyses of SR-depression and PHQ2 were 7.8% (z=28.74, sample prevalence 27.1%, population prevalence 20%) and 5.5% (z=14.0), respectively. Genetic correlation between depression phenotypes ranged between 0.59 and 1.21, with lower r_g_ identified between measures of depressive symptoms and case-control phenotypes (Figure 2a). Some of the genetic correlations from the LD score regression were greater than 1; genetic correlation from ldsc does not bound to 1, and the instances with values higher than 1 occurred when testing in the same sample with similar phenotype (rg 1.07, SE=0.0343) between ICD-Depression and SR-Depression within MVP), or between the somewhat smaller FinnGen sample and the large PGC/UKB broad depression (rg 1.21, SE=0.25) and 23andMe (rg=1.07, SE=0.21) samples. LD-intercept (1.03, SE 0.011) and attenuation ratio (0.0297, SE 0.011) of the LD score regression revealed minimal evidence for inflation or confounding.

**Figure 2.**
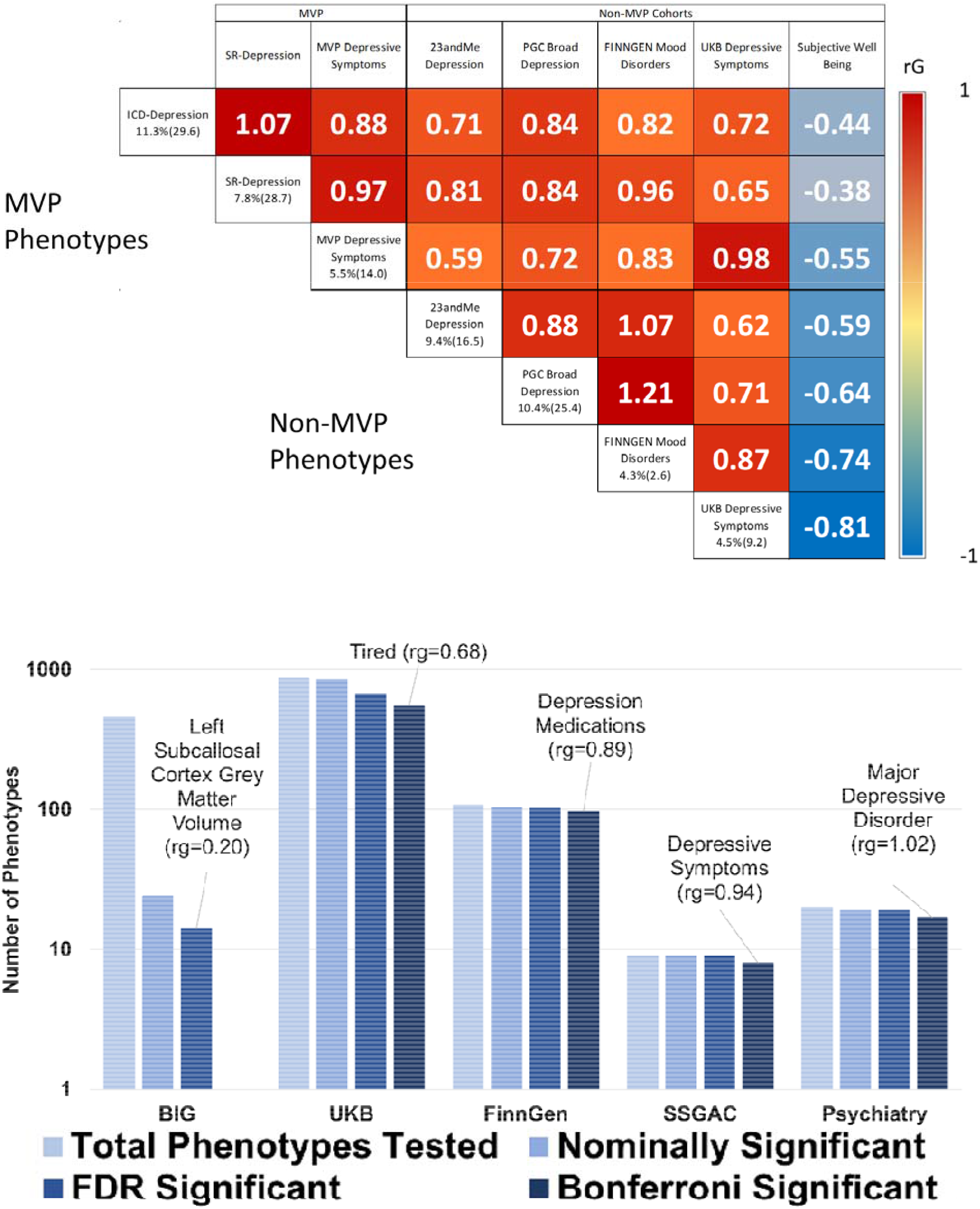
**Genetic Correlation.** Upper Panel. Genetic correlations between depression phenotypes, with subjective well-being included as a negative correlation comparator. Heritability (z-score) is given along the left axis of the matrix for each depression phenotype. Values within the matrix represent r_g_. Lower Panel. Summary of genetic correlation between ICD-Depression and 1,457 phenotypes from large-scale genetic studies of mental health and behavior. The Psychiatry category contains phenotypes from the Psychiatric Genomics Consortium, GWAS & Sequencing Consortium of Alcohol and Nicotine use, Million Veteran Program, and International Cannabis Consortium. The labels Tired and left subcallosal cortex grey matter volume represent UKB Field ID 2080 and BIG Field ID 0078, respectively.

Based on significant heritability estimates (h2 z>4), 1,457 traits from available GWAS summary statistics were sufficiently powered to assess genetic correlation with ICD-Depression. After multiple testing correction (p=0.05/1,457 trait pairs=3.43×10^−5^), 669 phenotypes were significantly genetically correlated with ICD-Depression (Figure 2b, Supplementary File 1). The most significant phenotypic correlations with ICD-Depression from each depressive trait category were: (i) depressive symptoms (SSGAC) r_g_=0.943–0.029, p=1.76×10^−228^, (ii) depression medications (FinnGen) r_g_=0.890–0.063, p=6.22×10^−45^, (iii) major depressive disorder (Psychiatry) r_g_=1.02±0.017, p<1.39×10^−300^ (note that LD score regression does not bound genetic correlation to 1),^12^ and (iv) frequency of tiredness/lethargy in last 2 weeks (UKB Field ID 2080) r_g_=0.684±0.018, p<1.39×10^−300^. No brain imaging phenotype met corrected significance criteria for genetic correlation with ICD-Depression; the most significantly genetically correlated brain imaging phenotype, using data provided from the Oxford Brain Imaging Genetics (BIG) project,^13^ relative to ICD-Depression was left subcallosal cortex grey matter volume (BIG Field ID 0078) r_g_=0.205±0.061, p=9.00×10^−4^.

### Transcriptome-Wide Association Study (TWAS)

Gene-based association analysis was performed by integrating GWAS association statistics and eQTL data of all brain and whole-blood tissues from GTEx v8. To prioritize target genes further, joint effects of gene expression correlation across tissues was leveraged using S-MultiXcan.^14^ 153 genes and their best representative tissues were below the Bonferroni corrected significance threshold (1.79e^−7^) for predicted gene expression in 14 tissues (Figure 3A; Supplementary file 2). Top genes for each tissue tested were: Amygdala (ZKSCAN4, p=1.65×10^−12^), anterior cingulate cortex (L3MBTL2, p=1.09×10^−14^), caudate (ZNF184, p=1.85×10^−9^), cerebellar hemisphere (PGBD1, p=1.67×10^−13^), cerebellum (ZSCAN9, p=8.4×10^−17^), cortex (TMEM161B, p=1.84×10^−12^), frontal cortex (FAM120A, p=3.25×10^−10^), hippocampus (ZSCAN12, p=1.14×10^−18^), hypothalamus (NEGR1, p=3.19×10^−25^), nucleus accumbens (DRD2, p=1.87×10^−20^), putamen (LIN28B-AS1, p=2.13×10^−12^), spinal cord c-1 (HIST1H1B, p=2.90×10^−18^), substantia nigra (RP11-318C24.2, p=2.41×10^−12^), and whole blood (ZNF165, p=4.01×10^−11^).

**Figure 3.**
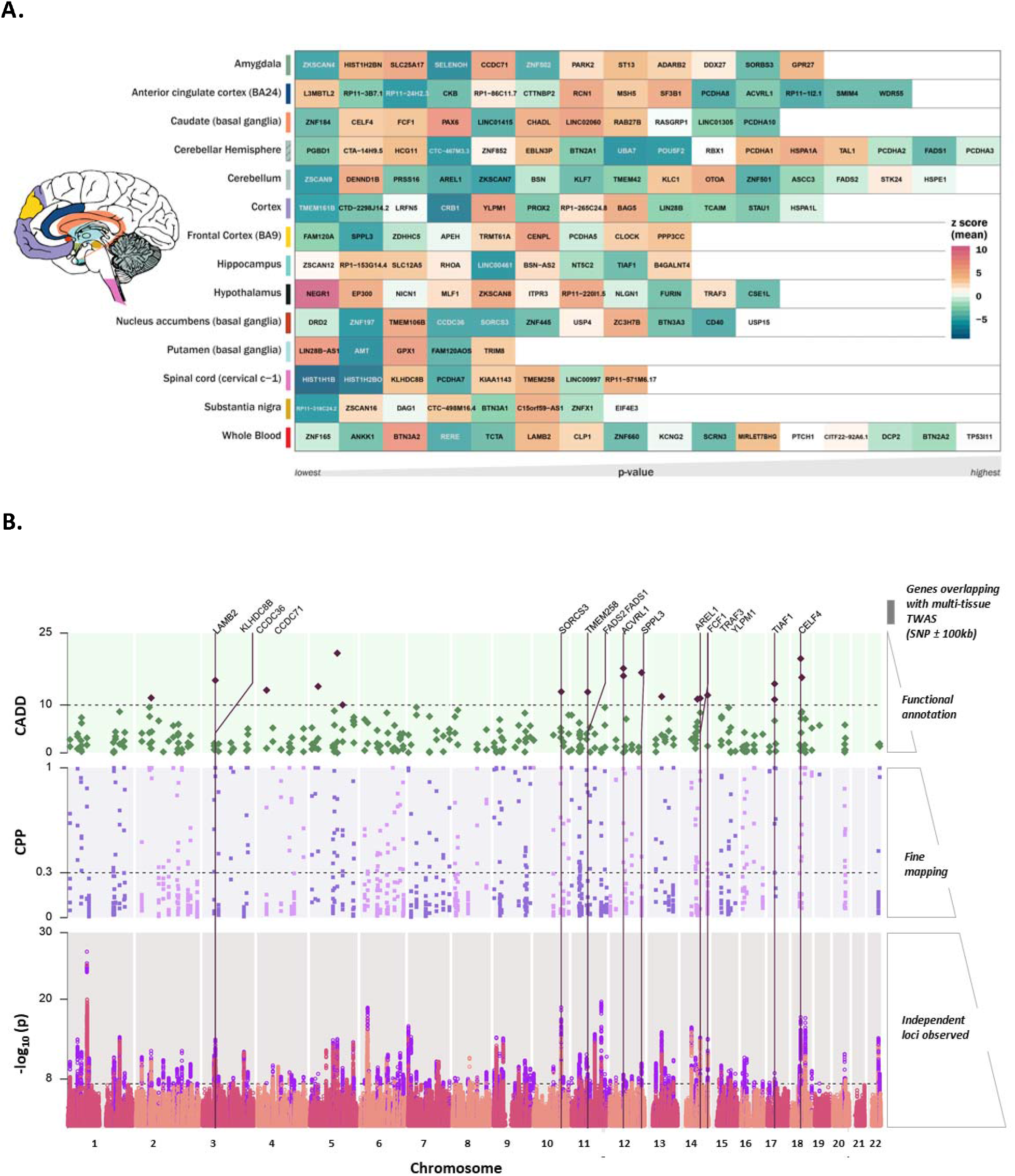
**A) Tissue-based gene association study (TWAS).** The genes were tested using MetaXcan for 13 brain tissues and whole blood from the GTEx-v8. The genes were compared across tissues to identify best representative tissues for each gene using SMultiXcan. Genes are arranged in order from left to right by respective tissue specific p-value, with the lowest value on the left. The color scale for the gene matrix is based on mean z-score. The values are reported in Supplementary file 2. **B) SNP prioritization using fine Mapping and functional scoring.** Bottom panel: Manhattan plot showing each genomic risk locus in violet. Middle panel: Each locus was fine mapped, and the causal posterior probability (CPP) on the y-axis is shown for SNPs from the causal set. The SNPs which had CPP ≥0.3 (30%) were annotated using Combined Annotation Dependent Depletion (CADD) scores. Top panel: The SNPs with CADD ≥ 10 are highlighted in purple; these SNPs were positionally mapped to 107 genes within 100kb. Only positional genes overlapping with multi-tissue TWAS results (Supplementary Figure 1) are annotated with vertical lines. Details of the prioritized SNPs are reported in Supplementary file

### Variant Prioritization

All 178 risk loci were fine-mapped (Figure 3B; bottom panel); 1620 SNPs in the causal set out of 14,016 GWS hits have higher posterior probability for causal relation with ICD-Depression (Figure 3B; middle panel). The SNPs with casual posterior probability ≥ 30% were annotated with Combined Annotation Dependent Depletion (CADD) score.^15^ There were 19 SNPs with CADD scores > 10, representing the top 1% of pathogenic variants across the human genome (Figure 3B; top panel). These SNPs were annotated to genes positioned within ±100kb. We found 17 genes overlapping with significant genes identified from cross-tissue TWAS analysis. Each gene-tissue pair was tested for colocalization of the region for eQTL and GWAS. The coloc^16^ method tests probability of four hypotheses (H_0–4_). Of these, H_4_ tests the hypothesis that the same locus is shared between GWAS and tissue-specific eQTL. Loci that were found to have 80% or higher probability for H_4_ were compared, to understand the LD structure and most prominent variant being shared by GWAS and eQTL. These gene-tissue pairs were CCDC71-Amygdala (H4-PP: 93.1%), FADS1-Cerebellar hemisphere (H4-PP: 96.6%), SPPL3-Frontal Cortex (H4-PP: 83.9%), TRAF3-Hypothalamus (H4-PP: 95.2%) and LAMB2-whole blood (H_4_-PP: 79.9%) (Supplementary file 2).

### Tissue expression analysis and genome-wide gene-based association study (GWGAS)

GWGAS conducted in MAGMA using the ICD-Depression GWAS meta-analysis identified 426 significant genes after Bonferroni correction for 16,038 protein coding genes. MAGMA tissue expression analysis identified enrichment across all brain tissues and pituitary using data from GTEX v8, with the strongest findings for Brodmann area 9 (p=7.31×10^−16^), and no enrichment in non-neuronal tissue (Figure S1).

### Gene Ontology

Gene ontology analysis conducted in ShinyGO^17^ identified 219 biological processes with FDR < 0.05, with top findings involved in nervous system development (q=1.20×10^−10^), synapse assembly (q=9.75×10^−9^), and organization (q=9.75×10^−9^) (Table S1).

### Drug mapping

The Manually Annotated Targets and Drugs Online Resource (MATADOR)^18^ database was tested for enrichment for 426 significant genes from the MAGMA analysis. This analysis identified 10 drug annotations with FDR < 0.05 including; diethylstilbestrol, Implanon, tamoxifen, raloxifene, nicotine, cocaine, cyclothiazide, felbamate, and riluzole.

### Latent Causal Variable

After filtering for suitable traits pairs with LCV-estimated h^2^ z-scores≥4, 1,667 phenotypes were powered to evaluate causal estimates relative to ICD-Depression; no statistically significant putatively causal genetic causality proportions (gĉps) were detected.

### Genomic structural equation modeling

(SEM) was used to evaluate how the ICD-Depression phenotype relates to 15 previously published large-scale GWAS of mental health and psychiatric phenotypes (See Online Methods and Discussion). Exploratory factor analysis (EFA) was conducted simultaneously on all traits and supported three- (cumulative variance=0.605) and four-factor models (cumulative variance=0.624) where each factor contributed over 10% to the cumulative explained variance. Anorexia nervosa did not load onto any factor during EFA and was therefore excluded from confirmatory factor analyses (CFA). CFA did not converge on a four-factor model due to high correlation between two factors. CFA of the three-factor model produced modest fit (comparative fit index=0.884; Figure 4, Supplementary File 3). Factor 1 generally represented internalizing phenotypes with major contributions from depressive symptoms(loading=0.95 ± 0.03), anxiety symptoms (loading=0.92 ± 0.03), and posttraumatic stress disorder (loading=0.92 ± 0.04). Factor 2 represented externalizing phenotypes with major contributions from risky behavior (loading=0.85 ± 0.03) and cannabis use disorder (loading=0.77 ± 0.04). Factor 3 represented educational attainment (loading=0.99 ± 0.03) and cognitive performance (loading=0.68 ± 0.03). ICD-Depression (DEP, Figure 4, Supplementary File 3) loaded onto factors 1 and, less strongly, on factor 2, independent of its covariance with all other phenotypes (DEP loading on Factor 1=0.77 ± 0.02; DEP loading on Factor 2=0.14 ± 0.02).

**Figure 4.**
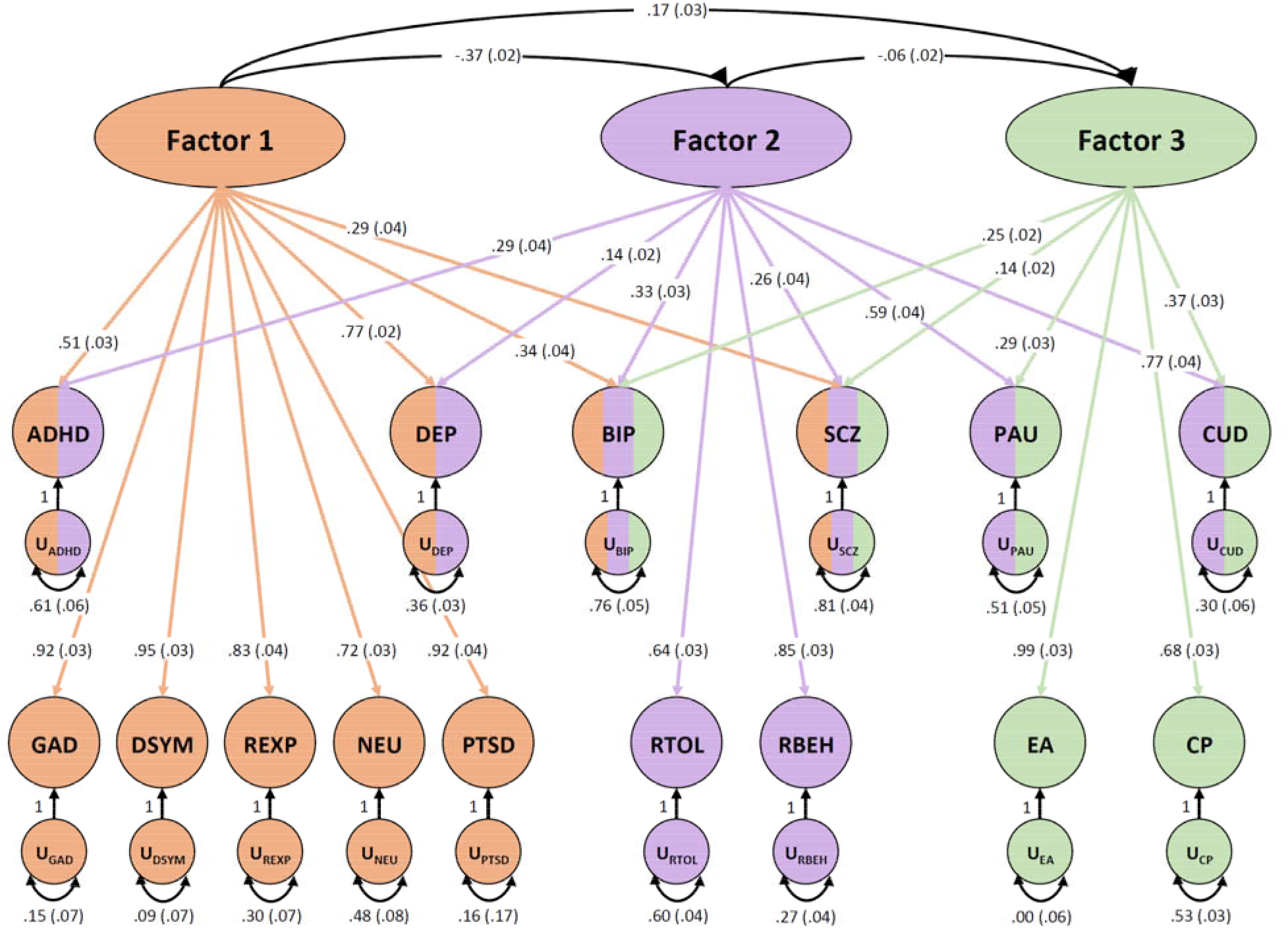
**Genomic SEM.** Genomic structural equation modeling of ICD-Depression meta-analysis (DEP) plus 14 additional traits. Exploratory factor analysis converged on a three-factor model. Arrows represent loading of each phenotype onto a connected factor with loading value and standard error provided for each. Multi-colored phenotypes indicate loading onto more than one factor while monochromatic phenotypes were unique to a single factor. Factors 1 generally represents internalizing symptoms, Factor 2 externalizing behaviors, and Factor 3, education/cognition. The correlation between factors is shown. Phenotype acronyms are: attention deficit hyperactivity disorder (ADHD), MVP ICD-Depression (DEP), bipolar disorder (BIP), schizophrenia (SCZ), problematic alcohol use (PAU), cannabis use disorder (CUD), generalized anxiety disorder (GAD), depressive symptoms (DSYM), reexperiencing (REXP), neuroticism (NEU), posttraumatic stress disorder (PTSD), risk tolerance (RTOL), risky behavior (RBEH), educational attainment (EA), and cognitive performance (CP).

### Ethics statement

The Central VA Institutional Review Board (IRB) and site-specific IRBs approved the MVP study. All relevant ethical regulations for work with human subjects were followed in the conduct of the study, and written informed consent was obtained from all participants.

## Discussion

We present the first genetic study of depression including more than a million informative participants, with new large analyses from the Million Veteran Program meta-analyzed with prior results from the PGC + UK Biobank, 23andMe, and FinnGen, the largest analysis so far in what is a fast-moving field. We investigated genetic correlation between three different definitions (ICD-Depression, SR-Depression, and PHQ2) of the depression phenotype within the MVP cohort. We identified 223 independently significant SNPs in 178 genomic loci associated with the primary meta-analysis, using an ICD code derived definition of depression for the MVP sample and GWAS summary statistics from 23andMe, UKB, PGC, and FinnGen. This is an improvement of 77 loci over the largest previous study that investigated a comparable phenotype.^10^ As these cohorts used somewhat different definitions for depression (Table1, Figure 1a, Methods), we also used LDSC to examine genetic correlations between MVP depression phenotypes and these differentially defined depression phenotypes in other outside independent cohorts. We investigated genetic correlation with 1,457 traits using available GWAS data, identifying 669 that were significantly correlated. We also used genomic structural equation modeling to evaluate how depression relates to other mental health and psychiatric phenotypes.

The MVP sample added substantially to our ability to discover new loci. Two of the most powerful prior studies conducted to date^7,8^ had substantial contributions from the UK Biobank. UK Biobank and MVP represent large and non-overlapping samples with consistent phenotypic assessments. This consistency in collection reduces ascertainment heterogeneity within samples and likely increases power to detect new loci. Adding another massive homogenously phenotyped sample here allowed us to discover 77 more loci than previously identified. It also provides a novel large independent cohort for conducting post-GWAS analyses, leveraging the substantial resources already produced by others in the field to improve understanding.

MVP is very informative for depression and related traits with several available measures, so we considered several different diagnosis definitions (Table 1), as follows. In the MVP, we considered (1) an ICD code-based algorithm to determine depression case status based upon diagnosis codes captured in the VA electronic health records (ICD-Depression), (2) self-reported diagnosis of depression as reported in the MVP Baseline Survey (SR-Depression), and (3) the 2 item PHQ scale of depressive symptoms in the past 2 weeks, included in the MVP Lifestyle Survey (depressive symptoms). Genetic correlations between these traits were high (r_g_ 0.81–1.07). We consider the first of these --ICD-Depression -- to be our “primary” analysis based on the larger explained heritability and sample size.

For meta-analyses of ICD-Depression and SR-Depression, we also used available GWAS summary statistics from 23andMe, UKB, PGC, and FinnGen (Table 1). Genetic correlation was conducted between the phenotypes to be meta-analyzed together to quantify potential heterogeneity between the studies to be combined. These studies used a variety of phenotype definitions, with some combining clinical diagnosis of depression based on structured interview and other broader methods,^7^ to self-reported treatment,^7^ to self-reported items on questionairres.^9^ This analysis is discussed in greater detail in the methods, but the genetic correlations between all traits ranged from 0.71–0.84.

The lead SNP from our primary analysis, rs7531118, (MAF=0.48, p=8.9×10^−29^) maps close to the neuronal growth regulator 1 gene (NEGR1) and is a brain eQTL for NEGR1. This SNP was at least nominally significant with concordant effect direction in all four studies included in this meta-analysis (MVP p=4.9×10^−5^, FinnGen p=0.04, PGC+UKB p=1.6×10^−17^, 23andMe p=2.8×10^−8^). The S-MultiXcan analysis prioritized hypothalamus as related to NEGR1. Recent work in Negr −/− mice have shown irregularities in several brain regions, including reduced brain volume in the hippocampus.^19^ Negr −/− mice showed abnormalities in social behavior and non-social interest. ^19^ Another study of Negr −/− mice identified a variety of depression-like and anxiety-like features in behavioral assays such as elevated plus maze and forced swim tests.^20^

The D_2_ dopamine receptor (DRD2) was another top finding from the TWAS analysis (Figure 3a), with significant predicted decreased expression in the nucleus accumbens. The mesolimbic dopamine reward circuit, of which nucleus accumbens is a critical part, has long been implicated in depression.^21^ A recent optogenetic study examining dopaminergic ventral tegmental area projections into nucleus accumbens found that dopamine receptors are required for the action of these neurons in depression-related escape behavior.^22^ Depression – like behavior in animals might be related to depression in humans through links to the reward system and symptoms of anhedonia. A recent randomized proof-of-mechanism trial^23^ investigated κ-opioid antagonists (KOR) as treatment for anhedonia symptoms. JNJ-67953964 was found to increase VTA activation relative to placebo during reward anticipation, highlighting the potential therapeutic mechanism by which KOR is thought to release inhibition on dopaminergic projections. The group receiving JNJ-67953964 showed reduced anhedonic symptoms relative to controls.^23^ That this gene and brain tissue emerged from hypothesis-free GWAS and TWAS tissue enrichment is a remarkable positive control with respect to known biology, and points to the potential value of other novel findings from this kind of research.

The CUGBP Elav-Like Family Member 4 (CELF4) gene has been highlighted recently in another depression GWAS study,^8^ and was our top finding for convergence between functional variant prioritization and multi-tissue TWAS results (Figure 3B, Supplemental Data Table 4). This gene is important in developmental disorders, with deletions of the 18q12.2 region which encompass the gene associated with autism spectrum disorder.^24,25^ Celf4 mutant mice show aberrations in sodium channel function, perhaps through increased NA_v_ 1.6 in the axon initial segment of excitatory neurons, and increased susceptibility to seizures.^26^ We agree with the assertion made in previous studies, now with additional functional and expression evidence, that CELF4 should be a focus of future brain research in depression and depression-like behaviors to elucidate its mechanism.

Genetic correlations with available GWAS summary statistics from 1,457 traits were conducted to assess overlap with other traits. There was high genetic correlation between our ICD-Depression meta-analysis and depression medication prescription in FinnGen (r_g_=0.89). This could be of value in evaluating depression phenotypes from large cohorts with access to linked electronic health records; anti-depressant medication prescription may be a viable proxy phenotype for depression diagnosis.

We used ShinyGO^17^ to identify overlap between top MAGMA genes and drugs of interest (Figure S2). Riluzole, an NMDA antagonist currently used to treat amyotrophic lateral sclerosis, was one of our top findings. This drug is currently in trials for combination therapy for treatment resistant depression.^27^ Another drug, cyclothiazide, is an allosteric modulator of AMPA (glutamatergic) receptors. Allosteric modulation of glutamatergic receptors has been considered a mechanistic treatment target for depression.^28^ This screen also identified an anti-seizure medication, felbamate, which has side effects including increasing depressive symptoms, suicidal ideation, and attempts. These enrichments, from hypothesis-free association with depression, show converging independent evidence from genetics of existing pharmacological targets based on underlying biological mechanisms.

Genomic SEM was used to investigate relationships between ICD-Depression and 15 other mental health and psychiatric phenotypes (Figure 4, Supplementary File 3). All traits tested except anorexia nervosa loaded onto at least one factor during exploratory analysis. Trait summary statistics come from the largest studies available. We identified three factors, with ICD-Depression loading onto the first two independently of covariance with the other phenotypes. Factor 1 may be thought to represent internalizing phenotypes, with major contributions from ICD-Depression, anxiety symptoms, and posttraumatic stress disorder. ICD-Depression also loaded (but less strongly) onto factor 2, which broadly represents externalizing phenotypes and psychosis, with the major contributions coming from risky behavior and cannabis use disorder. ICD-Depression did not load onto factor 3, which was mostly contributed to by educational attainment and cognitive performance and thus may represent a cognitive domain. Many of these GWAS studies, this one included, align themselves in ways consistent with existing theories of psychopathology, when clustered by genomic SEM.

We prioritized variants using biologically and statistically informed annotations. To prioritize genes and their target tissues we integrated both transcriptomics and CADD score prioritized variants. This method aided in the identification of shared causal loci for phenotype and tissue-specific eQTLs as evidenced by the high probability for 5 of the 17 genes tested. SNPs at CCDC71 (“Coiled-Coil Domain Containing 71”) have been reported to be associated with depressive symptoms in a multivariate genome wide association meta-analysis, and our prioritized SNP is in strong LD with their lead SNP (current study rs7617480, r2=0.83, D’=1.0).^29^ The FADS1 protein product, “Fatty Acid Desaturase 1” is involved in fatty-acid regulation and variants in this region have been reported to be associated with depression and substance use disorders. Treatment with omega-3 fatty acid may be beneficial in treatment of depression;^30^ though a role for omega-3 supplementation in treatment of depression is still controversial, and dietary omega-3 is probably insufficient to drive associations with (or treatment for) depression. There is nevertheless consistent evidence in the literature for an association with depleted omega-3 and increased depression risk.^30^ Variants in SPPL3, encoding “Signal Peptide Peptidase Like 3”, were reported to be associated with risk to major depression by Hyde and colleagues.^9^ The TRAF3 protein product, “TNF Receptor Associated Factor 3”, controls type-1 interferon response,^31^ and it has been reported that individuals treated with interferon are at high risk to develop depressive symptoms.^32^ LAMB2 is involved in neuropathic pain and influencing gene expression changes in brain pathways implicated in depression.^33^

Because no GWS findings were identified in our primary analysis of African ancestry we performed cross ancestry lookups in the summary statistics of European ancestry. Of 223 GWS SNPs from the European ancestry meta-analysis, 206 were available in African ancestry, 61% (n=125) had the same effect direction, 20 were nominally significant (p<0.05), and 1 SNP survived Bonferroni correction (Figure 5). This SNP that survived multiple testing correction (rs1950829 EUR p=7.24E×10^−19^, AFR p=9.34×10^−6^), is in an intron of the “Leucine rich repeat fibronectin type III domain containing 5” (LRFN5) gene. This gene was previously detected in genome-wide gene- and pathway based analyses of depressive symptom burden conducted in three cohorts from the Alzheimer’s Disease Neuroimaging Initiative (ADNI), the Health and Retirement Study (HRS), and the Indiana Memory and Aging Study (IMAS).^34^ As larger samples are collected for more diverse ancestry groups we expect to see more novel loci identified for non-European populations. Finally, we conducted a transancestry meta-analysis by combining studies of African and European ancestries in 1,213,867 participants, thereby identifying 233 independent SNPs and 183 risk loci. For now, trans-ancestral analysis is a way to leverage results from understudied populations.

**Figure 5.**
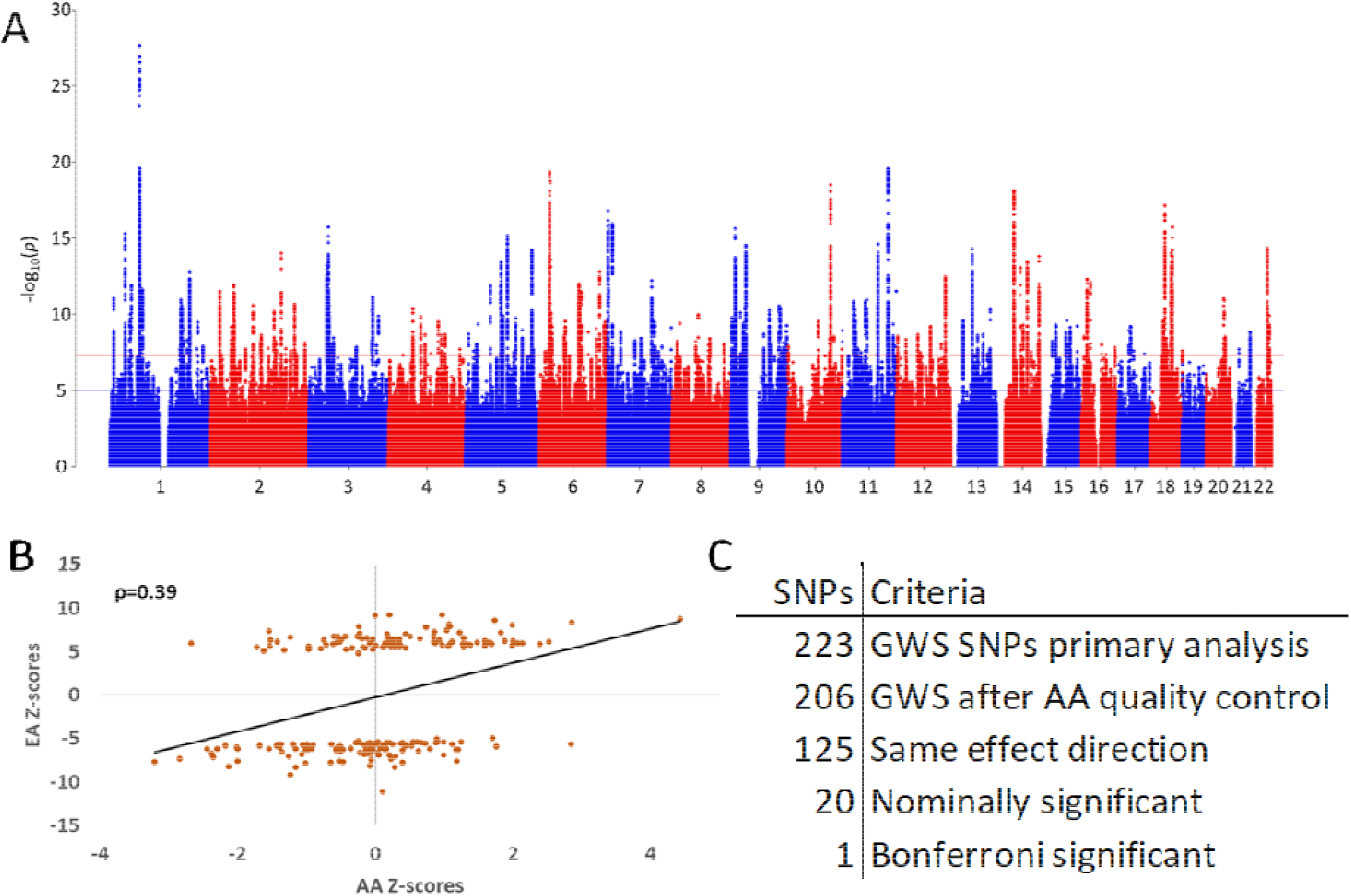
**Transancestry Meta-analysis.** A. Manhattan plot for transancestry meta-analysis of ICD-Depression (n=1,213,867). B. Scatter plot for 206 GWS SNPs (Spearman’s ?=0.39) from the primary ICD-Depression GWAS of different ancestries, plotting z-score for European ancestry (only) GWAS on the y axis and African ancestry (only) GWAS on the x axis. C. Overlap of SNPs from European and African ancestry GWASs. 223 GWS SNPs from the primary analysis, of which 206 are available in the AA GWAS following QC. 125 (61%) of the remaining SNPs had the same effect direction, 20 were nominally significant (p<0.05) and one was Bonferroni significant after correcting for 206 comparisons.

We recognize limitations in our study. Maximizing the power available for this analysis comes at the cost of accepting broader biobank phenotyping approaches, which may reduce specificity of findings for the core depression phenotype.^35^ Nonetheless, strong genetic correlations between the ICD derived depression with the broader definitions provide confidence in internal consistency, and future studies could look to further refine phenotyping. While all correlations were significant, there was substantial variance (95% CI=0.72–1.7) in correlations with the FinnGen sample, probably due to power and heterogeneity in the broad phenotype we used from this sample. Finally, other ancestries remain understudied in relation to Europeans. Perhaps the initial results reported here for the MVP African ancestry sample can help advance the field by encouraging additional concerted research in African and other non-European ancestral groups.

In summary, we identified substantially more loci than previous studies due to increased power, and several of these loci serve functions that should prioritize their further study in the pathology of major depression. We examined genetic correlations between depression GWAS other external phenotypes, largely confirming and strengthening previous observations. We showed substantial enrichments for several brain regions, such as hypothalamus and frontal cortex, known to be important for depression. We found overlapping biology with novel potential treatments using gene and drug-based enrichments. We used genomic structural equation modeling to show how the genetic architecture of depression fits into the context of other large GWAS of mental disorders and cognition, identifying emergent overlap from hypothesis-free GWAS approaches with existing theories of psychopathology with regard to clusters of internalizing and externalizing disorders.

## Online Methods

### Participants

The MVP cohort has been previously described.^36–39^ GWAS was conducted in each of two tranches of data separately by ancestry, depending upon when the data became available. Ancestry was assigned using 10 principal components (PCs) and the 1000 genomes project phase 3 EUR and AFR reference within each tranche of data. For the analysis of the quantitative phenotype we also performed a GWAS in the UK Biobank sample. Finally, we conducted GWAS meta-analyses of traits related to depression using data from 4 large cohorts (Table 1, Figure 1a): the Million Veteran Program (MVP),^34, 40^ the PGC/UK Biobank,^10^ FinnGen, and 23andMe.^9^ For the ICD definition of depression, the phenotype with the most available data for the MVP cohort, there were 1,154,267 total subjects for primary meta-analysis. For the secondary case control meta-analysis, we performed a similar analysis except we replaced the ICD-Depression diagnosis from MVP with the SR-Depression GWAS for a total of 1,057,768 participants. For the secondary analysis of depressive symptoms by PHQ, we included 286,821 total participants from UKB and MVP. We also performed a GWAS in the MVP African American (AA) sample of 59,600 participants. We included these participants in a trans-ancestral meta-analysis with a total sample size of 1,213,867 participants. Cohorts are detailed in Table 1.

### Phenotypes

Within MVP there were three depression phenotypes investigated across five different analyses. We used 1) an ICD code-based algorithm to determine depression case status based upon investigation of the electronic health records (ICD-Depression, primary analysis), 2) self-reported physician diagnosis of depression as reported in the MVP lifestyle survey (SR-Depression), and 3) the 2-item PHQ scale of depressive symptoms in the past 2 weeks, included in the MVP lifestyle survey (depressive symptoms). Phenotypes in outside cohorts for UKB-PGC and 23andMe have been previously described.^7–10^ See Table 1 and Figure 1 for summary. For the ICD code-based algorithm in MVP, codes used to assess case status are presented in Table S2. Cases included people with at least one inpatient diagnosis code or two outpatient diagnosis codes for Major Depressive Disorder (MDD). Controls include only those without any inpatient or outpatient depression diagnosis codes for depression.

### GWASs and meta-analyses

GWAS analysis was carried out in the MVP cohorts by logistic regression for ICD-Depression and SR-Depression and by linear regression for PHQ2 within each ancestry group and tranche using PLINK 2.0 on dosage data, covarying for age, sex, and the first 10 PCs. A similar GWAS was performed using linear regression in the UK Biobank samples, also using age, sex, and the first 10 PCs for PHQ2.

In individuals of European ancestry for ICD-Depression and SR-Depression, meta-analysis was performed using METAL with inverse variance weighting for: MVP tranche 1, MVP tranche 2, the PGC-UKB ICD-Depression meta-analysis,^10^ 23andMe,^9^ and FinnGen Mood [affective] disorders (http://r2.finngen.fi/pheno/F5_MOOD). For the PHQ2 meta-analysis, the procedures were the same for the following samples: MVP tranche 1, MVP tranche 2, and UK Biobank. Meta-analysis in the African American participants was carried out only between tranche 1 and 2 of the MVP data due to absence of data in the other samples.

The 23andMe phenotype was based on responses to 4 questions: “Have you ever been diagnosed by a doctor with any of the following psychiatric conditions?”, “Have you ever been diagnosed with clinical depression?”, “Have you ever been diagnosed with or treated for any of the following conditions? (Depression)”, and “In the last 2 years, have you been newly diagnosed with or started treatment for any of the following conditions? (Depression)”. Cases were defined as having responded “Yes” to any of the above questions, and controls, when not a case and at least 1 “No” response to the above questions.

The FinnGen diagnosis is defined by the F5 Mood category and was downloaded from Freeze 2 of the database (http://r2.finngen.fi/pheno/F5_MOOD). This phenotype is broad and contains manic episodes, bipolar disorders, depression, persistent mood disorders, and other unspecified mood [affective] disorders. Data from UKB^8^ is a broad depression phenotype based on affirmative responses to either of the questions: “Have you ever seen a general practitioner for nerves, anxiety, tension or depression?”, and “Have you ever seen a psychiatrist for nerves, anxiety, tension or depression?”. PGC data also has been previously reported,^7^ and come from meta-analysis of 35 cohorts with a spectrum of depression phenotypes, including some with clinical diagnosis from structured interviews and others with broader definitions.

## Post-GWAS analysis

### Linkage Disequilibrium Score Regression

For post-GWAS analysis, FinnGen was removed a priori due to potential for increased heterogeneity in the phenotype definition due to the broad nature of inclusion in the F5 Mood phenotype. Genetic correlation analyses were performed using LDSC to assess the degree of genetic overlap between phenotypes and across the cohorts included in the analysis. Per-trait observed-scale SNP-based heritability estimates were calculated via LDSC using the 1000 Genomes Project European linkage disequilibrium reference panel.^41,12^ Heritability estimates were calculated for 1,468 phenotypes from FinnGen, 4,083 phenotypes from UKB, 3,143 brain image derived phenotypes from the Oxford Brain Imaging Genetics (BIG) project, and phenotypes from the Psychiatric Genomics Consortium (PGC), the Social Science Genetic Association Consortium (SSGAC), and the Genetics of Personality Consortium (GPC). Heritability z-scores were calculated by dividing the heritability estimate per phenotype by its associated standard error. Phenotypes with heritability z-scores ≥ 4 were considered suitable for genetic correlation against ICD-Depression.^41,12^ For continuous UKB phenotypes we restricted our analyses to use inverse-rank normalized phenotypes instead of untransformed phenotypes. Genetic correlations are summarized by total phenotypes tested, nominally significant (p<0.05), and after application of 5% false discovery rate and Bonferroni thresholds (Figure 2b).

### Latent Causal Variable (LCV)

The LCV model was used to infer genetic causal relationships between trait pairs using the 1000 Genomes Project European linkage disequilibrium reference panel. ICD-Depression was subjected to LCV with all traits described above for genetic correlation analysis. Due to differences in heritability calculation method and the number of SNPs used by LCV versus LDSC, genetic correlation results were not used to inform LCV trait pair selection. Genetic causality proportions (gĉp) were interpreted only when the heritability zscore of both traits was ≥ 7, as determined by LCV, not LDSC.^42^ Fully causal relationships were deduced for significant trait pairs with gĉp estimates e 0.70; otherwise gĉp estimates were considered evidence for partial causality.^42^

### Genomic structural equation modeling (SEM)

Genomic SEM was performed using GWAS summary statistics in the genomicSEM and lavaan R packages.^43^ Exploratory factor analyses (EFA) were performed on 16 traits simultaneously (ICD-Depression [the main phenotype of interest for this study], attention deficit hyperactivity disorder, anorexia nervosa, bipolar disorder, cannabis use disorder, cognitive performance, depressive symptoms, educational attainment, anxiety symptoms, neuroticism, posttraumatic stress disorder, problematic alcohol use, reexperiencing, risk tolerance, risky behavior, and schizophrenia). EFAs were performed for 1 through N factors until the addition of factor N contributed less than 10% explained variance to the model. Confirmatory factor analysis was performed using the diagonally-weighted least squares estimator and a genetic covariance matrix of munged GWAS summary statistics for all 16 phenotypes based on the 1000 Genome Project Phase 3 European linkage disequilibrium reference panel.

### Transcriptome-Wide Association Study (TWAS)

We performed transcriptome-wide association study using MetaXcan for 13 brain tissues and whole blood using GTEx v8. The MetaXcan framework consists of two prediction models for GTEx v8; elastic net and MASHR-based model for deriving eQTL values. The MASHR model is biologically informed, with Deterministic Approximation of Posteriors (DAP-G) based fine mapped variables and recommended by the developers.^44^ Since the eQTL effect is shared across several tissues, the joint effect of eQTL in 14 tissues was tested using the S-MultiXcan, developed under the MetaXcan toolkit.^14^ We applied Bonferroni correction (corrected p-value threshold=1.79×10^−7^) for all gene-tissue pair tested.

### Variant prioritization

Each of the risk loci, determined from FUMA (default LD=0.6), were fine-mapped using CAVIAR.^45^ The set of causal SNPs were annotated with CADD^15^ scores followed by positional gene mapping within ±100kb. The genes that overlapped with significant gene cross-tissue eQTL analysis were further tested for colocalization. Coloc^16^ was used to test colocalization between specific gene eQTL tissue pairs (GTEx v8). The LocusCompareR R package was used to generate regional plots of tissue-specific eQTL and GWAS p-values.

### Genome-wide gene-based association study (GWGAS) and Enrichment Analysis

Summary statistics from the primary ICD-Depression meta-analysis were loaded into Functional Mapping and Annotation of Genome-Wide Association Studies (FUMA GWAS) to test for gene-level associations using Multi-Marker Analysis of GenoMic Annotation (MAGMA).^46^ Input SNPs were mapped to 17,927 protein coding genes. The GWS threshold for the gene-based test was therefore determined to be p=0.05/17,927=2.79×10^−6^. Genes from MAGMA’s gene-based association were used for gene ontology and drug-set enrichment using the ShinyGO^17^ web tool.

## Data Availability

Summary statistics will be made available in dbGAP by the Million Veteran Program following publication.

https://www.ncbi.nlm.nih.gov/projects/gap/cgi-bin/study.cgi?study_id=phs001672.v3.p1

## Supplementary Information

**Figure S1.**
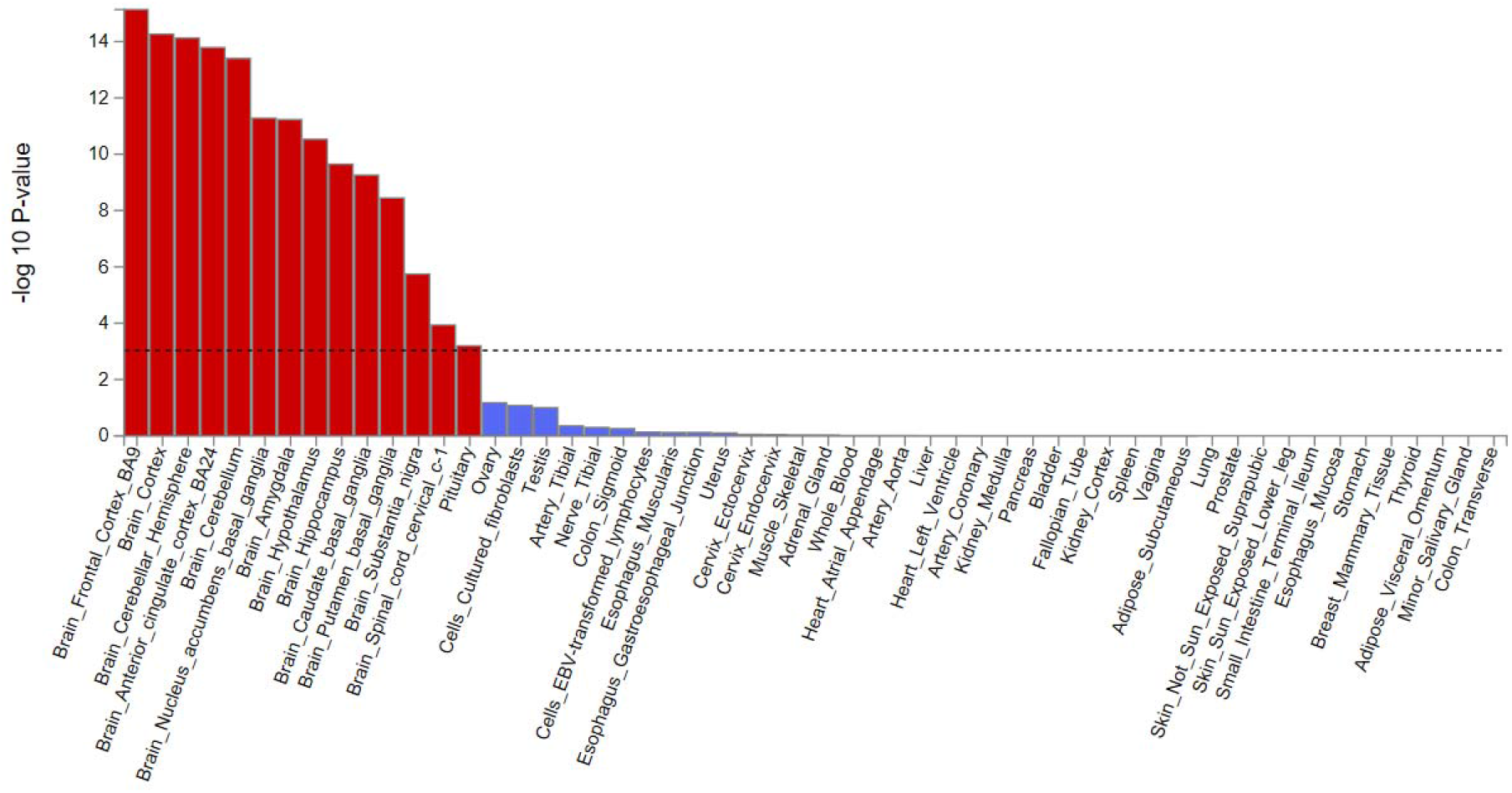
MAGMA Tissue Enrichment.

**Figure S2.**
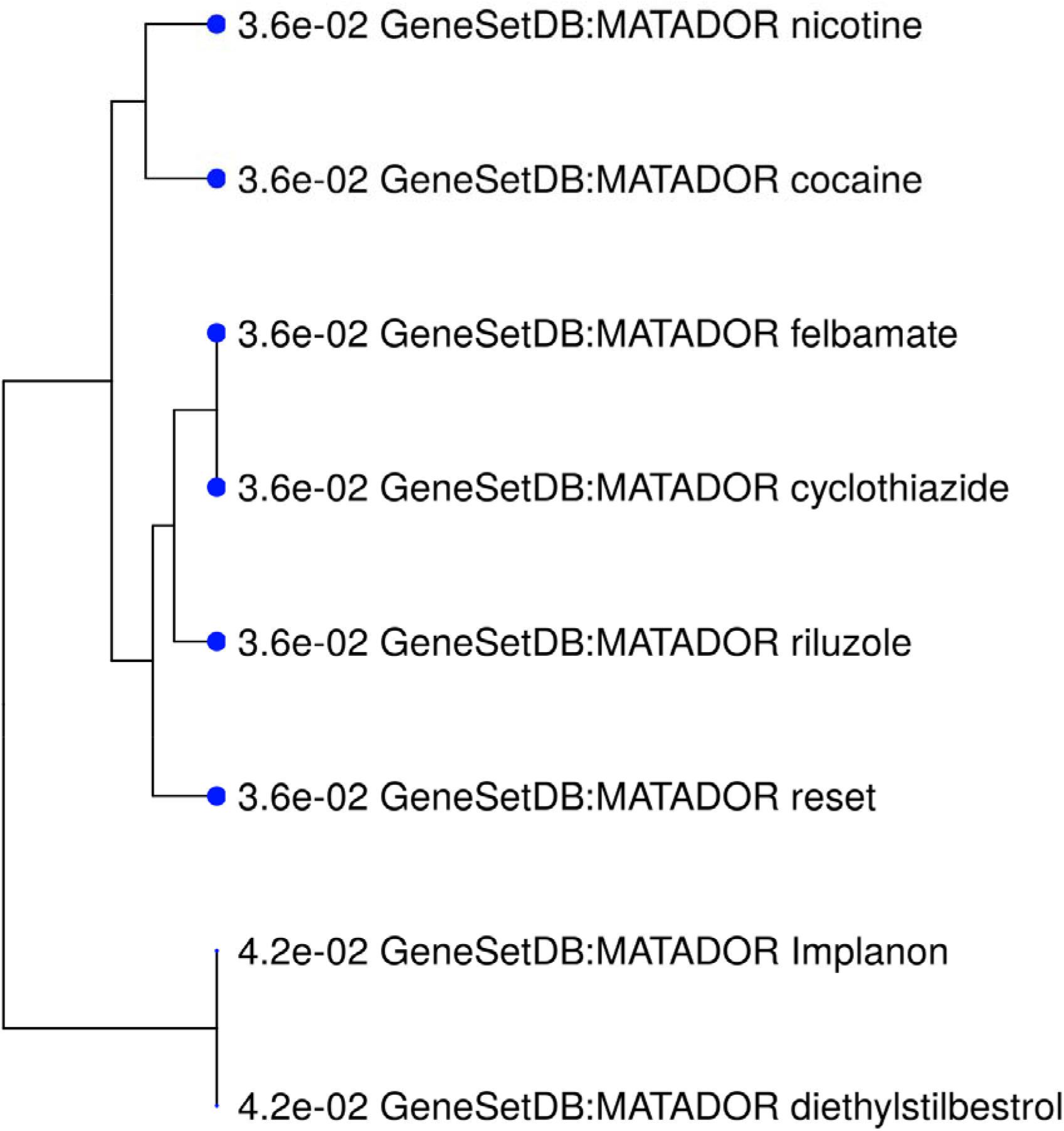
Drug repurposing.

**Table S1.** **Top Eight Gene Ontology Biological Processes. 219 Biological processes had an FDR < 0.05. The top eight processes with FDR < 1×10^−4^ are included here, the rest of the processes are reported in the supplemental data.**

| **Enrichment FDR** | **Genes in list** | **Total genes** | **Functional Category** |
| --- | --- | --- | --- |
| $1.20 \times 10^{-10}$ | 96 | 2474 | Nervous system development |
| $9.75 \times 10^{-9}$ | 21 | 182 | Synapse assembly |
| $9.75 \times 10^{-9}$ | 32 | 434 | Synapse organization |
| $2.74 \times 10^{-8}$ | 25 | 282 | Cell-cell adhesion via plasma-membrane adhesion molecules |
| $1.22 \times 10^{-7}$ | 19 | 172 | Homophilic cell adhesion via plasma membrane adhesion molecules |
| $1.15 \times 10^{-5}$ | 55 | 1412 | Neuron differentiation |
| $6.51 \times 10^{-5}$ | 57 | 1575 | Generation of neurons |
| $8.89 \times 10^{-5}$ | 35 | 762 | Synaptic signaling |

**Table S2.**
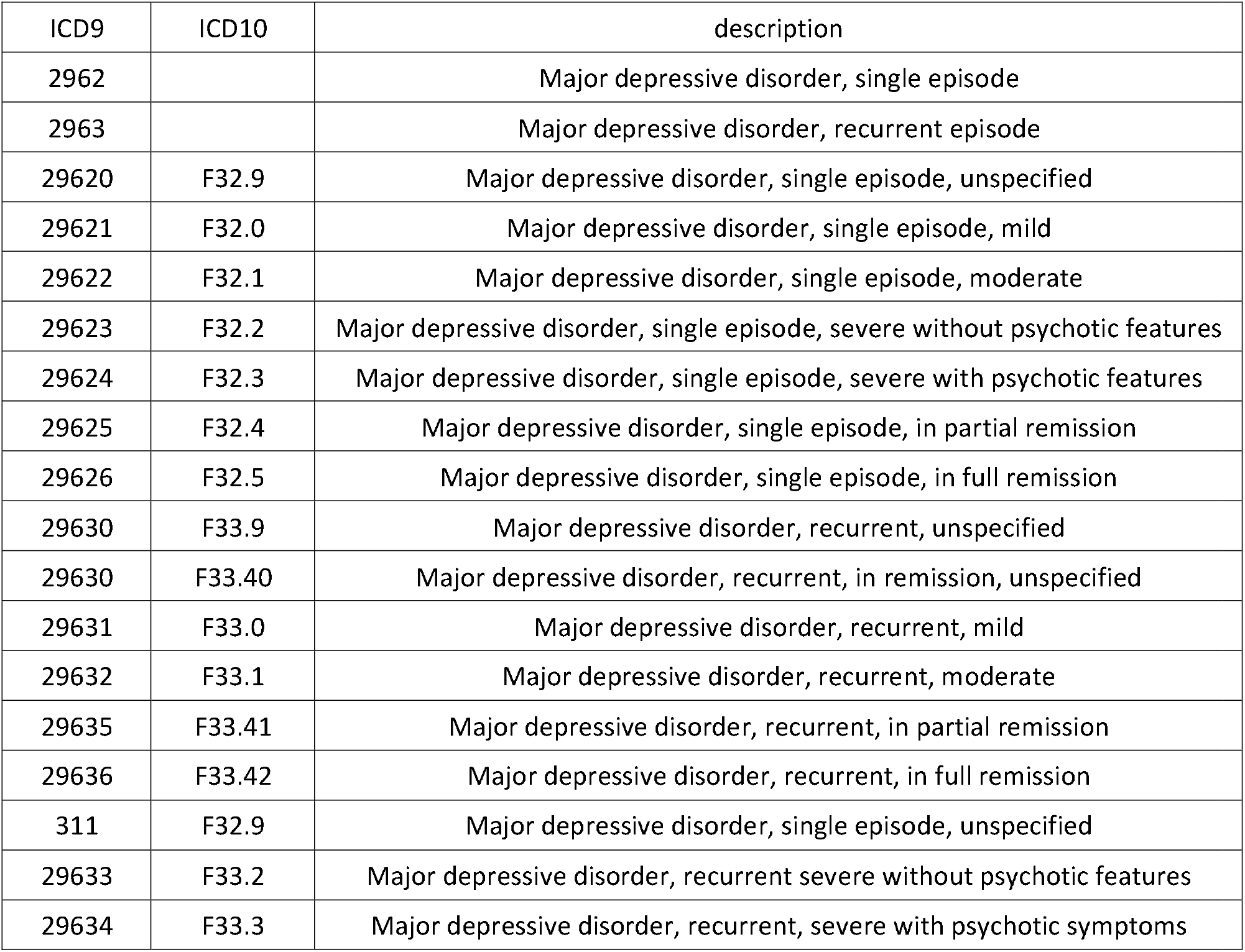
**ICD codes for MVP case status. Classification as a case required at least one inpatient code or two or more outpatient codes for Major Depressive Disorder (MDD). Classification as a control required neither no record of inpatient nor outpatient codes for MDD. Subjects with only one outpatient codes for MDD were excluded from all analyses.**

**Table S3.**
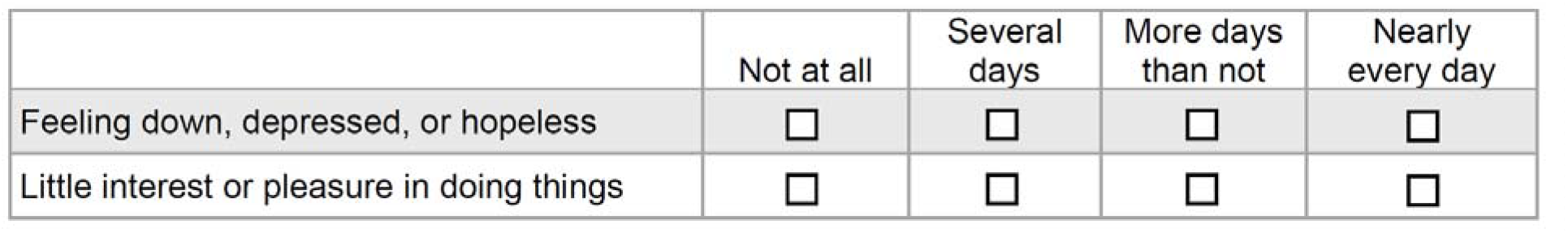
**PHQ-2 Phenotype, adapted from the MVP Lifestyle Survey**.

|  | Not at all | Several days | More days than not | Nearly every day |
| --- | --- | --- | --- | --- |
| Feeling down, depressed, or hopeless | ☐ | ☐ | ☐ | ☐ |
| Little interest or pleasure in doing things | ☐ | ☐ | ☐ | ☐ |

## Acknowledgements

We want to acknowledge the participants and investigators of the FinnGen study, 23andMe, the UK Biobank, PGC and the Million Veteran Program. We would like to thank the research participants and employees of 23andMe for making this work possible. We thank the veterans who participate in the Million Veteran Program.

## References

1. Hasin, D.S. et al. Epidemiology of Adult DSM-5 Major Depressive Disorder and Its Specifiers in the United States. JAMA Psychiatry 75, 336–346 (2018).

2. Roehrig, C. Mental Disorders Top The List Of The Most Costly Conditions In The United States: $201 Billion. Health Affairs 35, 1130–1135 (2016).

3. Pizzi, S.D. et al. Functional signature of conversion of patients with mild cognitive impairment. Neurobiology of Aging 74, 21–37 (2019).

4. Mullins, N. et al. GWAS of Suicide Attempt in Psychiatric Disorders and Association With Major Depression Polygenic Risk Scores. Am J Psychiatry 176, 651–660 (2019).

5. Strawbridge, R.J. et al. Identification of novel genome-wide associations for suicidality in UK Biobank, genetic correlation with psychiatric disorders and polygenic association with completed suicide. EBioMedicine 41, 517–525 (2019).

6. Levey, D.F. et al. Genetic associations with suicide attempt severity and genetic overlap with major depression. Transl Psychiatry 9, 22 (2019).

7. Wray, N.R. et al. Genome-wide association analyses identify 44 risk variants and refine the genetic architecture of major depression. Nat Genet 50, 668–681 (2018).

8. Howard, D.M. et al. Genome-wide association study of depression phenotypes in UK Biobank identifies variants in excitatory synaptic pathways. Nat Commun 9, 1470 (2018).

9. Hyde, C.L. et al. Identification of 15 genetic loci associated with risk of major depression in individuals of European descent. Nat Genet 48, 1031–6 (2016).

10. Howard, D.M. et al. Genome-wide meta-analysis of depression identifies 102 independent variants and highlights the importance of the prefrontal brain regions. Nat Neurosci 22, 343–352 (2019).

11. Kroenke, K., Spitzer, R.L. & Williams, J.B. The Patient Health Questionnaire-2: validity of a two-item depression screener. Med Care 41, 1284–92 (2003).

12. Bulik-Sullivan, B.K. et al. LD Score regression distinguishes confounding from polygenicity in genome-wide association studies. Nat Genet 47, 291–5 (2015).

13. Bycroft, C. et al. The UK Biobank resource with deep phenotyping and genomic data. Nature 562, 203–209 (2018).

14. Barbeira, A.N. et al. Integrating predicted transcriptome from multiple tissues improves association detection. PLoS Genet 15, e1007889 (2019).

15. Rentzsch, P., Witten, D., Cooper, G.M., Shendure, J. & Kircher, M. CADD: predicting the deleteriousness of variants throughout the human genome. Nucleic Acids Res 47, D886–D894 (2019).

16. Giambartolomei, C. et al. Bayesian test for colocalisation between pairs of genetic association studies using summary statistics. PLoS Genet 10, e1004383 (2014).

17. Ge, S.X., Jung, D. & Yao, R. ShinyGO: a graphical enrichment tool for animals and plants. Bioinformatics (2019).

18. Gunther, S. et al. SuperTarget and Matador: resources for exploring drug-target relationships. Nucleic Acids Res 36, D919–22 (2008).

19. Singh, K. et al. Neural cell adhesion molecule Negr1 deficiency in mouse results in structural brain endophenotypes and behavioral deviations related to psychiatric disorders. Sci Rep 9, 5457 (2019).

20. Noh, K. et al. Negr1 controls adult hippocampal neurogenesis and affective behaviors. Mol Psychiatry 24, 1189–1205 (2019).

21. Nestler, E.J. & Carlezon, W.A., Jr. The mesolimbic dopamine reward circuit in depression. Biol Psychiatry 59, 1151–9 (2006).

22. Tye, K.M. et al. Dopamine neurons modulate neural encoding and expression of depression-related behaviour. Nature 493, 537–541 (2013).

23. Krystal, A.D. et al. A randomized proof-of-mechanism trial applying the ‘fast-fail’ approach to evaluating kappa-opioid antagonism as a treatment for anhedonia. Nat Med (2020).

24. Barone, R. et al. Familial 18q12.2 deletion supports the role of RNA-binding protein CELF4 in autism spectrum disorders. Am J Med Genet A 173, 1649–1655 (2017).

25. Gilling, M. et al. A 3.2 Mb deletion on 18q12 in a patient with childhood autism and high-grade myopia. Eur J Hum Genet 16, 312–9 (2008).

26. Sun, W. et al. Aberrant sodium channel activity in the complex seizure disorder of Celf4 mutant mice. J Physiol 591, 241–55 (2013).

27. Sakurai, H. et al. Longer-term open-label study of adjunctive riluzole in treatment-resistant depression. J Affect Disord 258, 102–108 (2019).

28. Alt, A., Nisenbaum, E.S., Bleakman, D. & Witkin, J.M. A role for AMPA receptors in mood disorders. Biochem Pharmacol 71, 1273–88 (2006).

29. Baselmans, B.M.L. et al. Multivariate genome-wide analyses of the well-being spectrum. Nat Genet 51, 445–451 (2019).

30. Wani, A.L., Bhat, S.A. & Ara, A. Omega-3 fatty acids and the treatment of depression: a review of scientific evidence. Integr Med Res 4, 132–141 (2015).

31. Hacker, H., Tseng, P.H. & Karin, M. Expanding TRAF function: TRAF3 as a tri-faced immune regulator. Nat Rev Immunol 11, 457–68 (2011).

32. Chiu, W.C., Su, Y.P., Su, K.P. & Chen, P.C. Recurrence of depressive disorders after interferon-induced depression. Transl Psychiatry 7, e1026 (2017).

33. Descalzi, G. et al. Neuropathic pain promotes adaptive changes in gene expression in brain networks involved in stress and depression. Sci Signal 10(2017).

34. Nho, K. et al. Comprehensive gene-and pathway-based analysis of depressive symptoms in older adults. J Alzheimers Dis 45, 1197–206 (2015).

35. Cai, N. et al. Minimal phenotyping yields genome-wide association signals of low specificity for major depression. Nat Genet 52, 437–447 (2020).

36. Gelernter, J. et al. Genome-wide association study of post-traumatic stress disorder reexperiencing symptoms in > 165,000 US veterans. Nat Neurosci (2019).

37. Gelernter, J. et al. Genome-wide Association Study of Maximum Habitual Alcohol Intake in > 140,000 U.S. European and African American Veterans Yields Novel Risk Loci. Biol Psychiatry (2019).

38. Gaziano, J.M. et al. Million Veteran Program: A mega-biobank to study genetic influences on health and disease. J Clin Epidemiol 70, 214–23 (2016).

39. Harrington, K.M. et al. Gender Differences in Demographic and Health Characteristics of the Million Veteran Program Cohort. Womens Health Issues 29, S56–S66 (2019).

40. Levey, D.F. et al. Reproducible Genetic Risk Loci for Anxiety: Results From approximately 200,000 Participants in the Million Veteran Program. Am J Psychiatry, appiajp201919030256 (2020).

41. Bulik-Sullivan, B. et al. An atlas of genetic correlations across human diseases and traits. Nat Genet 47, 1236–41 (2015).

42. O’Connor, L.J. & Price, A.L. Distinguishing genetic correlation from causation across 52 diseases and complex traits. Nat Genet 50, 1728–1734 (2018).

43. Grotzinger, A.D. et al. Genomic structural equation modelling provides insights into the multivariate genetic architecture of complex traits. Nat Hum Behav 3, 513–525 (2019).

44. Barbeira, A.N. et al. Exploring the phenotypic consequences of tissue specific gene expression variation inferred from GWAS summary statistics. Nat Commun 9, 1825 (2018).

45. Hormozdiari, F., Kostem, E., Kang, E.Y., Pasaniuc, B. & Eskin, E. Identifying causal variants at loci with multiple signals of association. Genetics 198, 497–508 (2014).

46. Watanabe, K., Taskesen, E., van Bochoven, A. & Posthuma, D. Functional mapping and annotation of genetic associations with FUMA. Nat Commun 8, 1826 (2017).

